# Rapid quantitative electrochemical detection of SARS-CoV-2 antibodies in plasma and dried blood spot samples

**DOI:** 10.1101/2022.10.16.22281144

**Authors:** Sanjay S. Timilsina, Nolan Durr, Pawan Jolly, Donald E. Ingber

**Author notes:** Address all correspondence to: Donald E. Ingber, MD, PhD., Wyss Institute at Harvard University, CLSB5, 3 Blackfan Circle, Boston MA 02115 (. StataDX Inc., Boston, MA 02215, USA.

## Abstract

Coronavirus disease (COVID-19) caused by severe acute respiratory syndrome coronavirus 2 (SARS-CoV-2), which is a highly contagious disease with several variants, continues to spread as part of the global pandemic. With the roll-out of vaccines and development of new therapeutics that may be targeted to distinct viral molecules, there is a need to screen populations for viral antigen-specific SARS-CoV-2 antibodies. Here, we describe a rapid, multiplexed, electrochemical (EC) platform with on-chip control that enables detection of SARS-CoV-2 antibodies in less than 10 min using 1.5 µL of a patient sample. The EC biosensor demonstrated 100% sensitivity and specificity, and an area under the receiver operating characteristic curve of 1, when evaluated using 93 clinical samples, including plasma and dried blood spot samples from 54 SARS-CoV-2 positive and 39 negative patients. This EC biosensor platform enables simple, cost-effective, sensitive, and rapid detection of anti-SARS-CoV-2 antibodies in complex clinical samples, which is convenient for monitoring host humoral responses to vaccination or viral infection in broad population testing, including applications in low-resource settings. We also demonstrate the feasibility of using dried blood spot samples that can be collected locally and transported to distant clinical laboratories at ambient temperature for detection of anti-SARS-CoV-2 antibodies which can be used for serological surveillance and demonstrate the utility of remote sampling.

## 1. Introduction

The global pandemic caused by severe acute respiratory syndrome coronavirus 2 (SARS-CoV-2) has led to millions of infected individuals as well as global vaccination efforts.^[1]^ Real-time reverse transcription-polymerase chain reaction (RT-PCR) and rapid antigen detection are used to test for the presence of SARS-CoV-2 RNA and proteins in symptomatic and asymptomatic individuals.^[2]^ While this was extremely critical in the early phase of the pandemic, serologic testing of blood has become of greater interest more recently because it can detect the presence of antibodies against specific SARS-CoV-2 virus proteins.^[3]^ This is important because this can be used to assess whether patients had prior COVID-19 infection and determine the efficacy, duration, and longevity of vaccine responses, as well as qualify convalescent plasma for therapeutic purposes.^[4]^ Moreover, longitudinal evaluation of antibody titers in large populations is essential to determine the strength and duration of immunity generated by exposure to the primary virus, its variants, and vaccines, and all of this information is crucial for implementing effective public policy and vaccination strategies.^[5]^

Current SARS-CoV-2 vaccines induce the production of antibodies against SARS-CoV-2 Spike (S) protein, but not against its nucleocapsid (N) protein, while natural infection produces antibodies against both proteins, and thus diagnostic discrimination between these responses can help to differentiate natural immunity from vaccine-induced immunity.^[6]^ Traditional serological assays are not optimal for assessing antibody levels under pandemic conditions as enzyme-linked immunosorbent assays (ELISAs) take several hours to complete due to the need for multiple incubation and washing steps. Lateral flow immunoassays (LFIAs) are more rapid, but they produce less reliable results and are qualitative rather than quantitative.^[7]^ Similarly, while Rapid antigen detection tests (RADTs) now widely used in the United States for detection of viral proteins could be adapted for detection of antibodies, they have low sensitivity.^[8]^ Another option would be use of chemiluminescent enzyme immunoassays (CLIAs) (Abbott and Roche Elecsys), however, they are expensive, lack scalability, and require specific integrated instrument platforms.^[9]^ Many of these tests also must be carried out in a clinical research laboratory. Guidelines implemented to prevent spread of viral infections such as SARS-CoV-2 during serological surveillance also make collection of venous blood logistically difficult and have generated interest in dried blood spot (DBS) sampling. This is because the blood can be self-collected via a finger prick at-home or by community members and mailed to test sites at ambient temperatures.^[10]^ Through their ease of storage and shipment, low cost, and minimally invasive nature DBSs can be employed to screen large populations for antibody titers even in low-resource settings. DBSs only require a few microliters of blood, can be easily multiplexed and automated, and are compatible with a range of bioanalytical methods, namely chromatography, spectroscopy, and immunoassays.^[11]^ Also, DBS do not require refrigeration for shipment or storage and results obtained with these samples have been shown to correlate well with those obtained from plasma or serum using ELISAs or commercial SARS-CoV-2 antigen and antibody assays.^[12]^ DBS samples are also considered to be nonregulated and exempt materials for shipping if they are properly packaged making it easy to return DBS specimens from home directly to a clinical laboratory for testing.^[12b, 13]^ Thus, there is high demand for rapid and cost-effective detection approaches to monitor levels of different SARS-CoV-2 antibodies to assess patient responses infections and vaccines in a timely manner with high sensitivity and specificity, which may be used either at the point-of-care (POC) or in clinical laboratories in combination with use of DBS sampling.

Here, we describe an electrochemical (EC) biosensor platform that enables rapid, sensitive, and highly specific detection of anti-SARS-CoV-2 antibodies from both plasma and DBS in minutes (**Fig. 1**). The biosensor is coated with a previously reported antifouling nanocomposite coating composed of bovine serum albumin (BSA) cross-linked with glutaraldehyde (GA) and doped with highly conducting pentaamine-modified graphene nanoflakes (prGOx), which efficiently prevents biofouling and reduces non-specific binding thereby greatly increasing assay sensitivity. To develop a sensitive and specific serologic assay for COVID-19, specific ligands were covalently immobilized on the coated EC sensor to capture SARS-CoV-2 antibodies. The EC sensors also can be multiplexed to detect multiple different SARS-CoV-2 antibodies simultaneously while also including on-chip controls. Analysis of plasma and DBS samples from 93 SARS-CoV-2 patients revealed that the EC sensor platform displayed 100% sensitivity and 100% specificity with Area under the ROC Curve (AUC) of 1 in less than 10 min using only 1.5 µL of sample.

**Figure 1.**
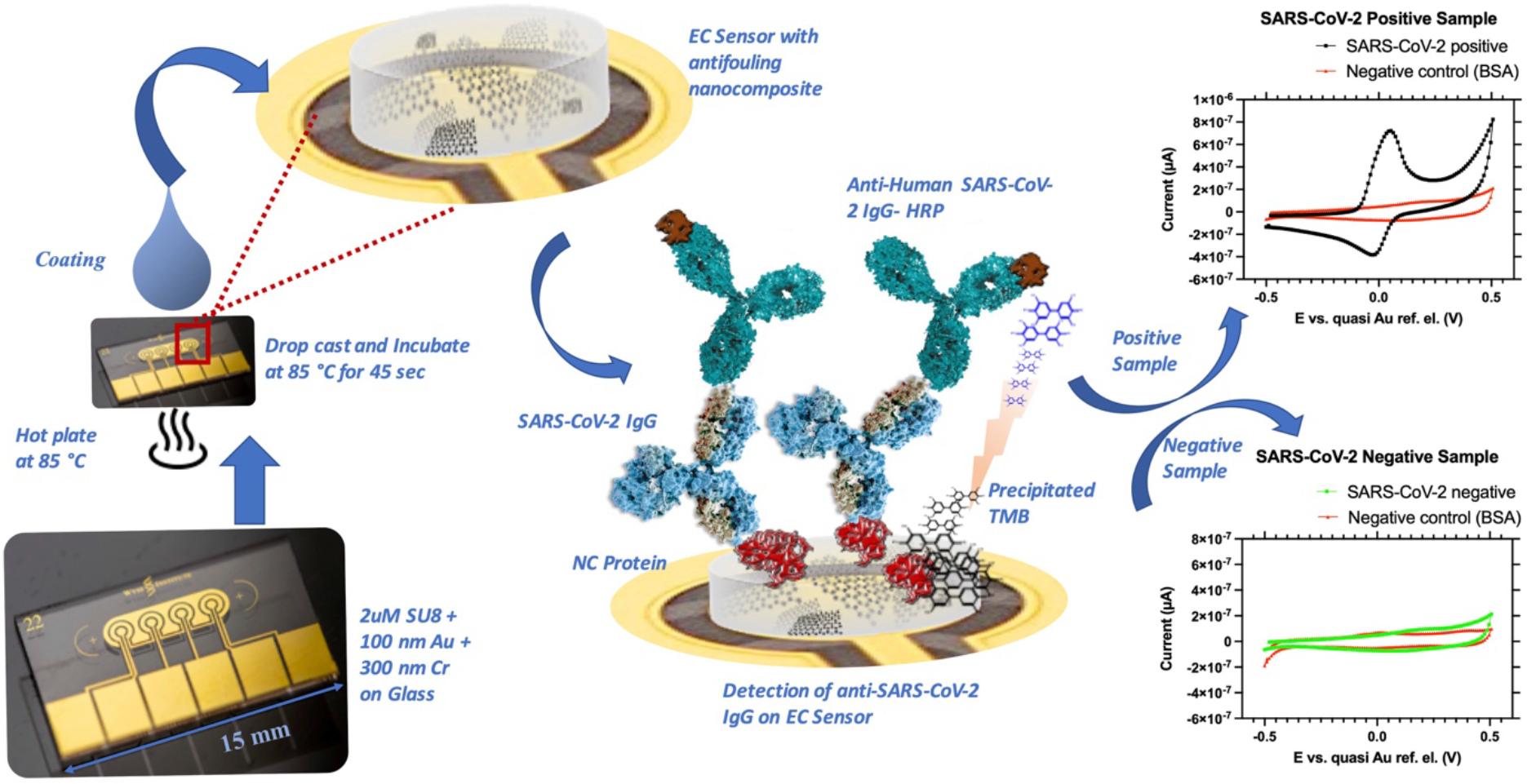
Schematic of the multiplexed EC sensor platform. The antifouling nanocomposite is rapidly drop cast on the gold electrodes of the EC sensor (left). A sandwich assay was used to measure SARS-CoV-2 IgG bound to Nucleocapsid (NC Protein) that was pre-immobilized on the nanocomposite coating above sensor where TMB precipitates to generate an EC signal (middle). The top and bottom graphs represent CV for positive and negative SARS-CoV-2 samples, respectively, with on-chip negative control.

## 2. Methods

### 2.1 Fabrication of Electrochemical (EC) sensor

EC sensor with gold electrodes purchased from Telic Company were custom fabricated using a standard photolithography process. The gold sensor electrode chips were cleaned as described previously.^[14]^ The BSA/prGOx/GA anti-fouling nanocomposite was prepared by mixing 8 mg/mL of prGOx (tetraethylene pentamine functionalized reduced graphene oxide) (Millipore Sigma, no. 806579) with 5 mg/mL BSA (IgG-Free, Protease-Free, Bovine Serum Albumin) (Jackson ImmunoResearch, no. 001-000-162) or Exbumin (Recombinant human albumin excipient, InVitria, no. 777HSA097) in 10 mM phosphate-buffered saline solution (PBS, pH 7.4) (Sigma Aldrich, USA, no. D8537). The solution was mixed using a vortex mixer and sonicated in a tip sonicator for 30 min using 1 s on/off cycles at 50% amplitude, 125 W and 20 kHz (Bransonic, CPX 3800), followed by heating (Labnet, no. D1200) at 105 ^0^C for 5 min to denature the protein. The mixture was kept at 4 ºC until further use. Before applying antifouling nanocomposite to the EC-sensor, the resulting opaque black mixture was centrifuged at 16.1 relative centrifugal force for 15 min to remove the excess aggregates. The semi-transparent nanocomposite supernatant solution was then mixed with 70% glutaraldehyde (Sigma Aldrich, USA, no. G7776) for crosslinking in the ratio of 70:2. Cleaned and plasma-treated (0.5 mbar and 50% power for 8 min) sensors were kept over a hot plate for 2 min for the temperature to equilibrate to 85 ºC. 70 µL of anti-fouling nanocomposite was then drop cast to each chip and incubated for 45s.^[15]^ The sensors were immediately washed by dipping in PBS, rinsed at 400 rpm for 10 min, and dried with a slide spinner (Millipore Sigma, no. 674664).

### 2.2 Detection of SARS-CoV-2 IgG in EC-Biosensor and ELISA

1-ethyl-3-(3-dimethyl aminopropyl) carbodiimide hydrochloride (EDC, Thermo Fisher Scientific, no. 22980) and N-hydroxysuccinimide (NHS, Sigma Aldrich, no. 130672) were used to conjugate the capture protein to the surface of the sensor with antifouling nanocomposite. Briefly, EDC (400 mM) and NHS (200 mM) were dissolved in 50 mM MES (2-(N-morpholino) ethanesulfonic acid) buffer (pH 6.2) and deposited on the gold sensor electrodes with antifouling nanocomposite for 30 min at room temperature in the dark for surface activation. Sensors were then quickly rinsed with Milli Q water and dried with compressed air before spotting of capture protein (0.6 mg/mL) on top of the three working electrode areas using Xtend capillary microarray Pin (LabNEXT, no. 007-350). The fourth working electrode was always spotted with 5 mg/mL BSA/recombinant Human Albumin (rHA) (InVitria, no. 777HSA097) as a negative control and stored overnight at 4 ºC in a humidity chamber. After conjugation, biosensors were washed with PBS and quenched with 15 µL of 1M ethanolamine (SigmaAldrich, no. E9508) for 30 min and blocked with 10 µL of 2.5% BSA/rHA in PBS for 1 hour.

Detection of anti-SARS-CoV-2 IgG was performed on the biosensor using the optimized conditions. Three working electrodes were spotted with a capture N protein (SARS-CoV-2 Nucleocapsid protein (His Tag), GenScript, no. Z03480) diluted in PBS (0.6 mg/mL). SARS-CoV-2 positive or negative plasma samples were diluted 10-fold using 1 % BSA/rHA in PBS. Mouse monoclonal anti-Human IgG Fc (abcam, no. ab99759) linked with HRP (horseradish peroxidase) was diluted to 40 µg/mL in 1 % BSA/rHA in PBS. Diluted clinical samples were mixed with detection antibody in the ratio of 9:1 and 15 µL was added to each biosensor and incubated for seven minutes. Biosensors were then washed with PBST (PBS with 0.05 % Tween 20 (Sigma Aldrich, no. P9416)). 10 µL of precipitating 3,3′,5,5′-Tetramethylbenzidine (TMB, Sigma-Aldrich, USA, no. T9455) was then added to biosensors and incubated for 1 min before washing. Finally, measurement was performed in 10 µL of PBST using a potentiostat (Autolab PGSTAT128N, Metrohm) by cycling the potential between -0.5 and 0.5 V with a scan rate of 1 V/s *vs*. on-chip integrated gold quasi-reference electrode. Peak height was then calculated using Nova 1.11 software for data analysis.

For standard ELISA of the biomarkers, 100 µL of 1 µg/mL capture N protein in carbonate-bicarbonate buffer at pH 9.2 was added to Nunc™ MaxiSorp™ ELISA plates (BioLegend, no. 423501) and incubated overnight at 4 ºC. The plates were washed 3 times with 200 µL of PBST followed by the addition of 200 µL of 5% Blotto (Fisher Scientific, no. NC9544655) for 1h. After washing the plates, 100 µL of 200-fold diluted clinical plasma samples were added and incubated for 30 min. After rewashing the plates, 100 µL of 50 ng/mL of horseradish peroxidase (HRP)-linked detection antibody was added for 30 min. The plate was then washed and 150 µL of turbo TMB (Thermo Scientific, no. 34022) was added for 20 min, followed by 150 µL of Stop solution to stop the reaction. The plate was immediately read using a microplate reader at 450 nm. All clinical samples were collected under the approval of the Institutional Review Board for Harvard Human Research Protection Program (IRB21-0024).

### 2.3 Screening of detection antibody and capture Nucleocapsid (N) protein

For screening of detection antibody, N protein from Raybiotech was immobilized on EC-biosensor and ELISA. For ELISA, 8 positive and 8 negative and for EC-biosensor, 4 positive and 2 negative SARS-CoV-2 samples were tested. Six screened detection antibodies include Goat anti-Human IgG Fc Secondary Antibody, HRP (ThermoFisher Scientific, no. A18817); Goat anti-Human IgG (Fc specific)−Peroxidase (Millipore Sigma, no. A0170); Peroxidase AffiniPure F(ab’)_2_ Fragment Goat anti-Human IgG, Fcγ (Jackson ImmunoResearch, no. 109-036-170); Mouse monoclonal anti-Human IgG Fc-HRP (abcam, no. ab99759); Human IgG-Fc Fragment Antibody (Bethyl Laboratories, no. A80-148P); and AffiniPure Goat anti-human IgG, Fc fragment specific (Jackson ImmunoResearch, no. 109-005-008). Similarly, for screening of capture N protein 8 positive and 6 negative (ELISA) and 7 positive and 5 negative (EC-biosensor) SARS-CoV-2 samples were tested. 5 different N proteins screened include, SARS-CoV-2 Nucleocapsid protein (GenScript, no. Z03480); Recombinant SARS-CoV-2 Nucleocapsid Protein (RayBiotech, no. 230-30164); SARS-CoV-2 Nucleocapsid (AA 1-419) protein (antibodies-online, no. ABIN6952315); SARS-CoV-2 Nucleocapsid-His recombinant Protein (SinoBiological, no. 40588-V08B); and SARS-CoV-2 Nucleoprotein, His-Tag (NativeAntigen no. REC31851).

### 2.4 Assay Development and optimization

Sandwich ELISA was performed both on EC-sensor and ELISA. Initially, a feasibility study was performed where a biotin-labeled detection antibody followed by the addition of Poly-HRP-Streptavidin (Thermo Fisher Scientific, no. N200) was replaced by HRP labeled detection antibody to reduce the assay steps and complexity. The assay was performed on both ELISA and EC-sensor with 2 positive, 2 negative, and blotto as a negative control. An assay for the optimization of the concentration of detection antibody was then performed on EC-biosensor. 4 positive and 3 negative SARS-CoV-2 samples were tested with different concentrations of detection antibody ranging from 5-80 µg/mL. Likewise, optimization of the concentration of capture N protein from 0.1-0.9 µg/mL was performed using 2 positive and 3 negative SARS-CoV-2 samples.

Similarly, the optimum sample dilution study was performed using undiluted and diluted samples (5 - 15-fold in 1% BSA/rHA) with 3 positive and 2 negative SARS-CoV-2 samples. Finally, the optimum assay time was studied where different incubation time of the sample (7-12 min) was performed against TMB incubation time of 1 and 2 min. Furthermore, a titration study was carried out to validate the clinical sample dilution and incubation timing. Three high titer positive samples and two negative samples were taken for the titration study and clinical samples were diluted from 0 - 1000-fold before performing the assay. In addition, a study was performed to compare the effectiveness of rHA over BSA. The assay was performed with 5 positive and 4 negative SARS-CoV-2 samples along with BSA/rHA as a negative control. To overcome the regulatory hurdle of using BSA in a medical device, nanocomposite coating was prepared with BSA and rHA, which was evaluated by performing the whole EC assay. Briefly, the coating was prepared by replacing BSA with rHA followed by functionalization with 0.6 mg/mL N protein or 5mg/mL BSA/rHA as a negative control. Then, the chips were blocked with 15µL of ethanolamine followed by 10µL of 2.5% BSA/rHA and used the respective buffer to run 10-fold diluted SARS-CoV-2 samples (5 positive and 4 negative).

### 2.5 Anti-SARS-CoV-2 IgG detection on EC-sensor using Clinical samples

Optimized assay conditions were used to run clinical samples on EC sensors. 23 SARS-CoV-2 positive and 21 SARS-CoV-2 negative clinical samples were purchased from Ray Biotech and BWH Crimson Core Laboratory and kept at -80 °Cuntil further use. An additional 23 positive and 14 negative SARS-CoV-2 samples were purchased from National Institute for Biological Standards and Control (NIBSC). Similarly, 8 SARS-CoV-2 positive and 4 SARS-CoV-2 negative Dried Blood Spots on Whatman filter paper #903 were purchased from RayBiotech (CoV-DBS-1) and kept at -80 °Cuntil further use. For reconstitution of DBS, Whatman filter paper with dried blood was kept in a tube and 300 µL of 0.05% PBST was added and kept on a shaker overnight. The reconstituted DBS was then directly used for the assay of anti-SARS-CoV-2 IgG. A time-point study of SARS-CoV-2 was also carried out to understand seroconversion better. Plasma samples were collected on 5 different days (10-21) after the onset of symptoms of COVID-19 for each person. The study was performed with 3 different patients. To run anti-N SARS-CoV-2 Rabbit IgG assay, we immobilized the EC sensor with 0.6 mg/mL N protein or 5mg/mL rHA as a negative control. After blocking the chip, 15 µL of different concentrations of SARS-CoV-2 rabbit IgG samples were spiked to pre-pandemic plasma samples and incubated on the chip for 30 min. Then, the chips were washed and 10 µL of 1:1000 Anti-rabbit IgG-HRP (RayBiotech, no. 130-10760-100) prepared in 1% rHA was added to the chip for 10 min. Chips were washed, and finally, precipitating TMB was added for 1 min before washing and EC measurement. For the calibration curve of NIBSC diagnostic calibrant, the assay was run using the optimized condition in 1% rHA and pre-pandemic plasma sample.

## 3. Results

### 3.1 Single-Step Assay Development

A sandwich ELISA was performed using the EC sensor and an ELISA during assay development. An initial feasibility study was performed where a biotin-labeled detection antibody (followed by washing and addition of streptavidin-polyHRP) was replaced by HRP-labeled detection antibody (single-step assay) to reduce the assay steps and complexity. Using traditional ELISA, with both biotin and HRP-labeled detection antibody, a significantly higher signal for positive samples was observed compared to negative SARS-CoV-2 samples for both the viral N and S proteins (**Fig. S1a**,**b)**. However, on the EC platform, the HRP-labeled detection antibody showed a higher signal for positive samples and no signal for negative samples as compared to the EC assay with biotin detection antibody where a small signal for 1 negative sample was observed **(Fig S1c**,**d)**. Thus, the HRP-linked detection antibody was used for further experiments because it significantly reduces complexity and assay time without compromising assay sensitivity. After the feasibility of the HRP-labeled detection antibody was established, the anti-SARS-CoV-2 IgG detection antibody concentration was varied from 5-80 µg/mL to establish a single-step assay (**Fig. S2)**. The concentration-dependent increases in positive signals were observed and 40 µg/mL of detection antibody was determined to be the optimum concentration, which was used in all further experiments.

### 3.2 Screening of detection antibodies for capture of N protein

Studies have shown that the sensitivity and specificity for the serological assay of SARS-CoV-2 antibody is significantly affected by choice of capture antigen and detection antibody (human anti-SARS-CoV-2).^[16]^ We screened six commercially available SARS-CoV-2 IgG detection antibodies to find the antibody with the highest sensitivity and specificity for the EC platform. Using ELISA, all the detection antibodies showed a similar pattern for positive and negative samples, where up to 25% of positive samples displayed lower signals than one of the negative samples. However, the Sigma and Abcam detection antibodies had comparatively lower signals for negative controls **(Fig. S3)**. When the EC biosensor was used, detection antibodies obtained from Sigma (**Fig. 2a**), Jackson (**Fig. 2b**), and Invitrogen (**Fig. 2c**) could not clearly differentiate between positive and negative samples. However, with Abcam detection antibody, even the low titer positive sample gave a significantly higher signal than negative samples (**Fig. 2d**). Thus, as the Abcam detection antibody could differentiate all positive and negative samples, it was used for further experiments to obtain a highly sensitive and specific assay.

**Figure 2.**
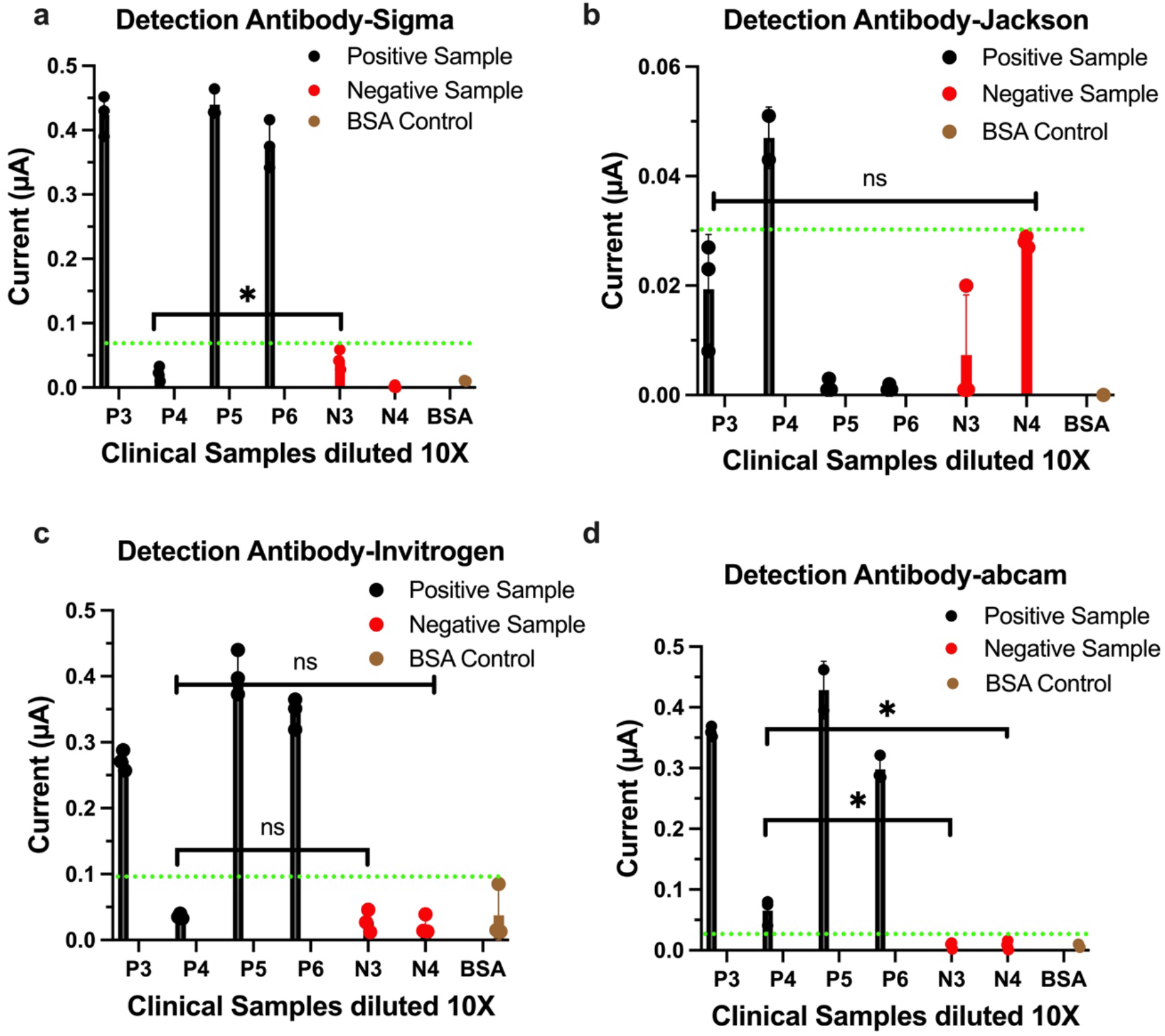
Screening of anti-human SARS-CoV-2 IgG detection antibody on EC-biosensor from four commercial sources, including Sigma (a), Jackson Immunoresearch (b), abcam (c), and Invitrogen (d). Error bars represent the s.d. of the mean; n=3; P= SARS-CoV-2 positive sample; N=SARS-CoV-2 negative sample; and BSA=Negative Control. Green dotted line represents highest signal for negative sample/control. Statistical analysis was performed by unpaired t-test (ns P > 0.05; *P < 0.05); all two-tailed.

All viral proteins elicit antibody responses to some extent, but it is necessary to identify proteins to which the immune system best responds and produces the highest affinity antibodies to increase the sensitivity and specificity of a serological assay. Moreover, the more unique the protein is, the lower the cross-reactivity with other coronaviruses.^[17]^ Thus, we then screened various sources of N protein for their ability to be detected by the Abcam antibody. When tested by ELISA, use of the Genescript N protein resulted in one negative sample showing a similar signal as a positive sample and when N protein from Sinobiological and AIC was used, some positive samples showed lower signals than negative one (**Fig. S4b**,**c)**. With other N capture proteins, 3 or more positive samples showed lower signals than some negative samples (**Fig. S4**). When we screened N proteins in the EC biosensor, the N protein from Sinobiological (**Fig. 3a**), AIC (**Fig. 3b**), Native Antigen Company (**Fig. 3c**) produced aberrant signals that did not scale with positive or negative samples. Finally, N capture protein obtained from Raybiotech and Genscript both generated signals for all positive samples, however, the Genscript protein had lower signals in the negative samples (**Fig. 3d,e**). Thus, Genscript N protein was used for further experiments because it showed high sensitivity and specificity and could differentiate all positive and negative controls.

**Figure 3.**
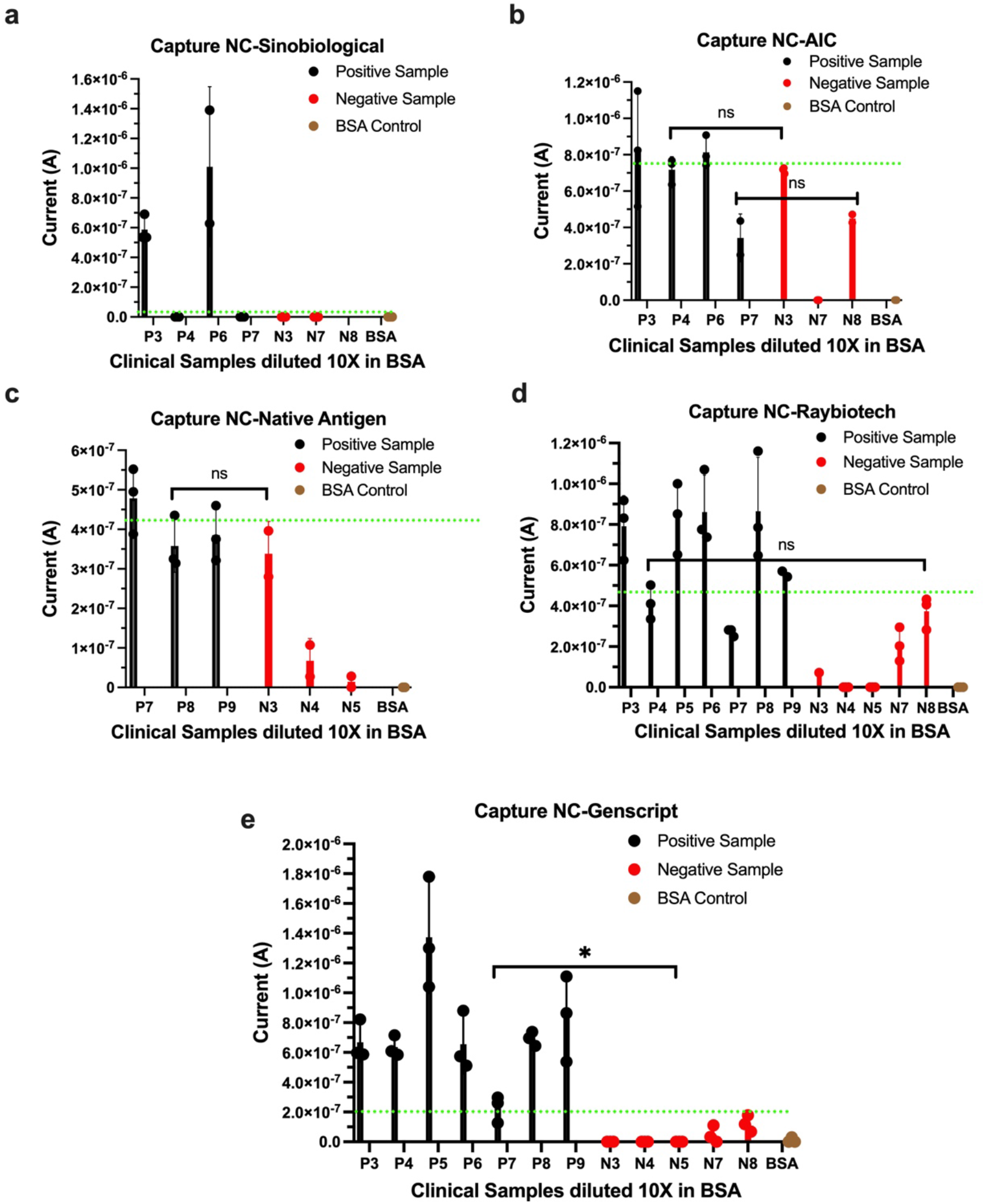
Screening of SARS-CoV-2 N capture protein on EC-biosensor from five commercial sources, including Raybiotech (a), Sinobiological (b), Advanced ImmunoChemical (c), Native Antigen Company (d), and Genscript (e). Error bars represent the s.d. of the mean; n=3; P= SARS-CoV-2 positive sample; N=SARS-CoV-2 negative sample; and BSA=Negative Control. Green dotted line represents highest signal for negative sample/control. Statistical analysis was performed by unpaired t-test (ns P > 0.05; *P < 0.05); all two-tailed. Green dotted line represents signal for negative sample with the highest value.

### 3.3 Assay optimization

The coating density of N protein on the electrode surface is critical to achieve the optimal surface-to-volume ratio necessary for efficiently capturing and detecting anti-SARS-CoV-2 IgG in the test sample. Thus, to reduce non-specific binding and increase assay sensitivity, we examined the effect of varying N protein concentration (from 0.1 to 0.9 µg/mL). These studies revealed the optimum concentration of capture N protein to be 0.6 µg/mL (**Fig. 4a**).

**Figure 4.**
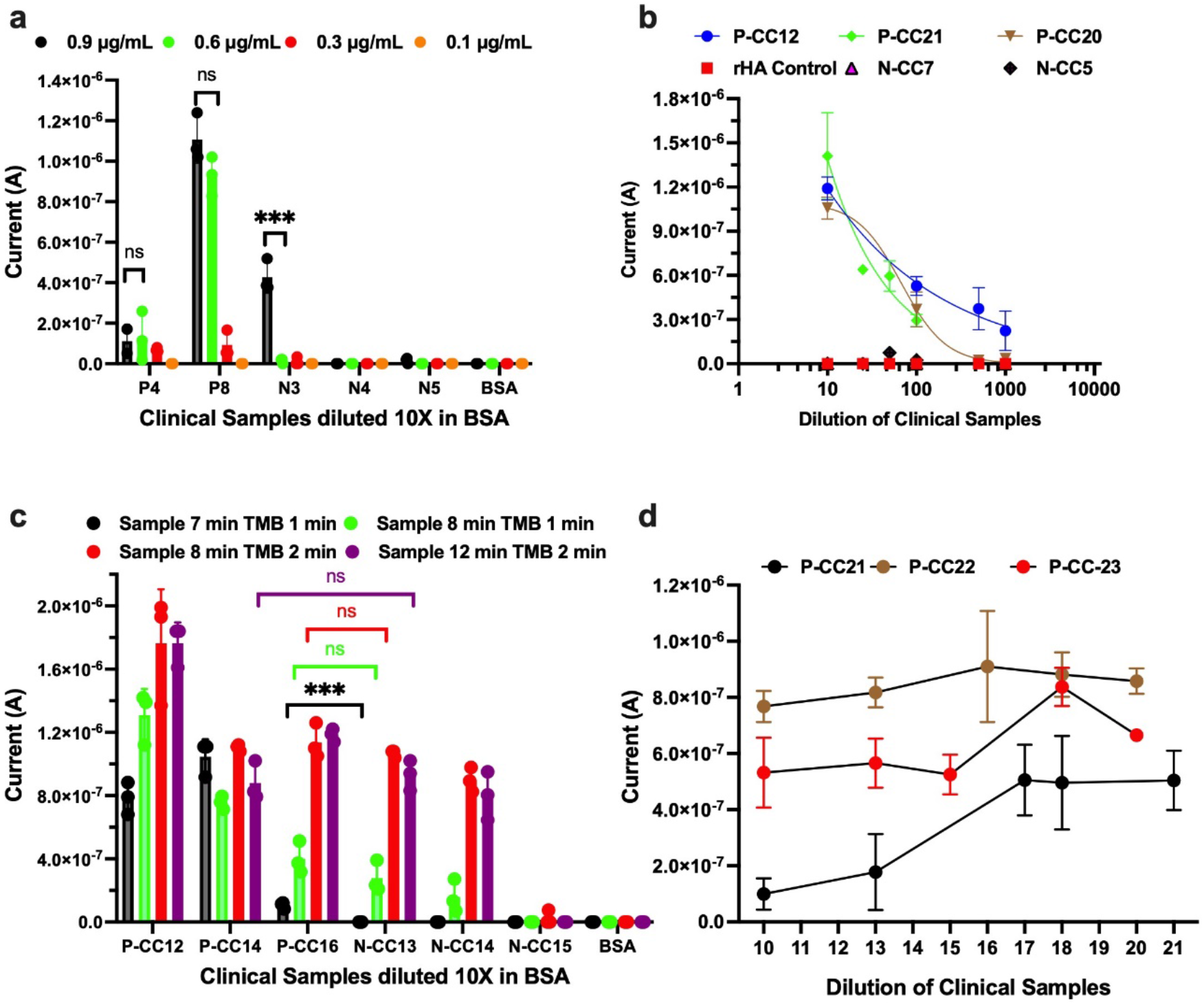
Assay development and optimization on EC-sensor for detection of SARS-CoV-2 IgG. (a) Optimization of concentration of capture N protein. (b) Titration of high titer SARS-CoV-2 positive clinical samples. (c) Optimization of sample and TMB incubation time for rapid detection of SARS-CoV-2 IgG. (d) Time-point study of anti-SARS-CoV-2 IgG clinical sample from 10-21 days after the onset of symptoms. Error bars represent the s.d. of the mean; n=3; P and P-CC= SARS-CoV-2 positive sample; N and N-CC=SARS-CoV-2 negative sample; and BSA=Negative Control. Statistical analysis was performed by unpaired t-test (ns P > 0.05; *P < 0.05; **P < 0.01; ***P < 0.001); all two-tailed.

To avoid the hook and matrix effect, we studied the effect of sample dilution (0 to 15-fold) for the detection of anti-SARS-CoV-2 IgG. In general, an increase in signal was observed with positive samples from undiluted to 5- and 10-fold dilution, saturating at 10- to 15-fold (**Fig. S5a**). Thus 10-fold dilution was considered optimum because below this level of dilution the signal was suppressed, likely due to combined hook and matrix effect.

To better understand the dilution required and the feasibility of rapid testing as a quantitative method, a titration study was performed where high titer anti-SARS-CoV-2 IgG clinical samples were serially diluted from 0 to 1000-fold (**Fig. 4b** and **Fig. S5b**). Signal generated with positive samples increased with 0- to 10-fold dilution, possibly due to the hook’s effect. As expected, the negative samples did not show any signal, and the signal for all positive samples decreased proportionally to the increase in dilution from 10- to 1000-fold. Thus, the titration study also revealed that a 10-fold dilution of the sample is optimal for the study.

To perform rapid detection of anti-SARS-CoV-2 IgG with high sensitivity and specificity, we also optimized the sample incubation time and TMB concentration. With sample incubation time of 7 min and TMB time of 1 min (black bar), all the positive samples showed signals while all negative samples did not (**Fig. 4c**). When sample incubation time was increased to 8 or 12 min with a TMB time of 1 or 2 min, we lost the ability to discriminate between some positive and negative samples. Thus, a sample incubation time of 7 min and TMB precipitation time of 1 min was considered to the optimum. In addition, to understand the seroconversion, we performed a time point study using an anti-SARS-CoV-2 IgG clinical sample in the three patient groups. As expected, the levels of anti-SARS-CoV-2 IgG generally increased over the first 17 days from the onset of symptoms which is consistent with previous reports (**Fig. 4d**).^[18]^

We also performed an assay to observe if BSA in the antifouling coating and the blocking buffer could be replaced by recombinant human albumin (rHA) without compromising sensitivity and specificity of the assay. We explored rHA as it is blood and animal component-free recombinant human albumin excipient which is U.S. Food and Drug Administration (FDA) and (European Medicines Agency) EMA-approved excipient.^[19]^ rHA Both BSA and rHA-based coating produced similar signals for all positive samples, however, there was less signal in the negative samples using rHA (**Fig. S6**).

### 3.4 Validation of EC-Biosensor using Clinical Samples

We then used the EC sensor to test 46 positive and 35 negative SARS-CoV-2 clinical plasma samples to validate its usefulness. These studies revealed that all negative samples showed minimum or no signal and all the positive samples showed a significantly higher signal (**Fig. 5a**). Analysis of the ROC curve revealed that the assay displayed 100% sensitivity and 100% specificity with AUC=1 **(Fig. 5b)**. We then carried out the same assay using 12 DBS samples, which confirmed that the EC sensor can be used to detect IgG in reconstituted dried blood without any further treatment. All positive DBS samples showed signal while none of the negative samples did, which again shows 100% sensitivity and specificity (**Fig. 5 c,d)**. The high sensitivity and specificity of the EC sensor may be attributed to the highly efficient antifouling BSA/prGOx/GA coating, which has very low non-specific binding and allow us to perform the assay in plasma with minimum matrix effect. For a proof-of-concept quantitative assay, the anti-N SARS-CoV-2 rabbit IgG calibration curve was run on the EC sensor and we detected a sensitivity of 1 ng/mL (**Fig. S7a)**.

**Figure 5.**
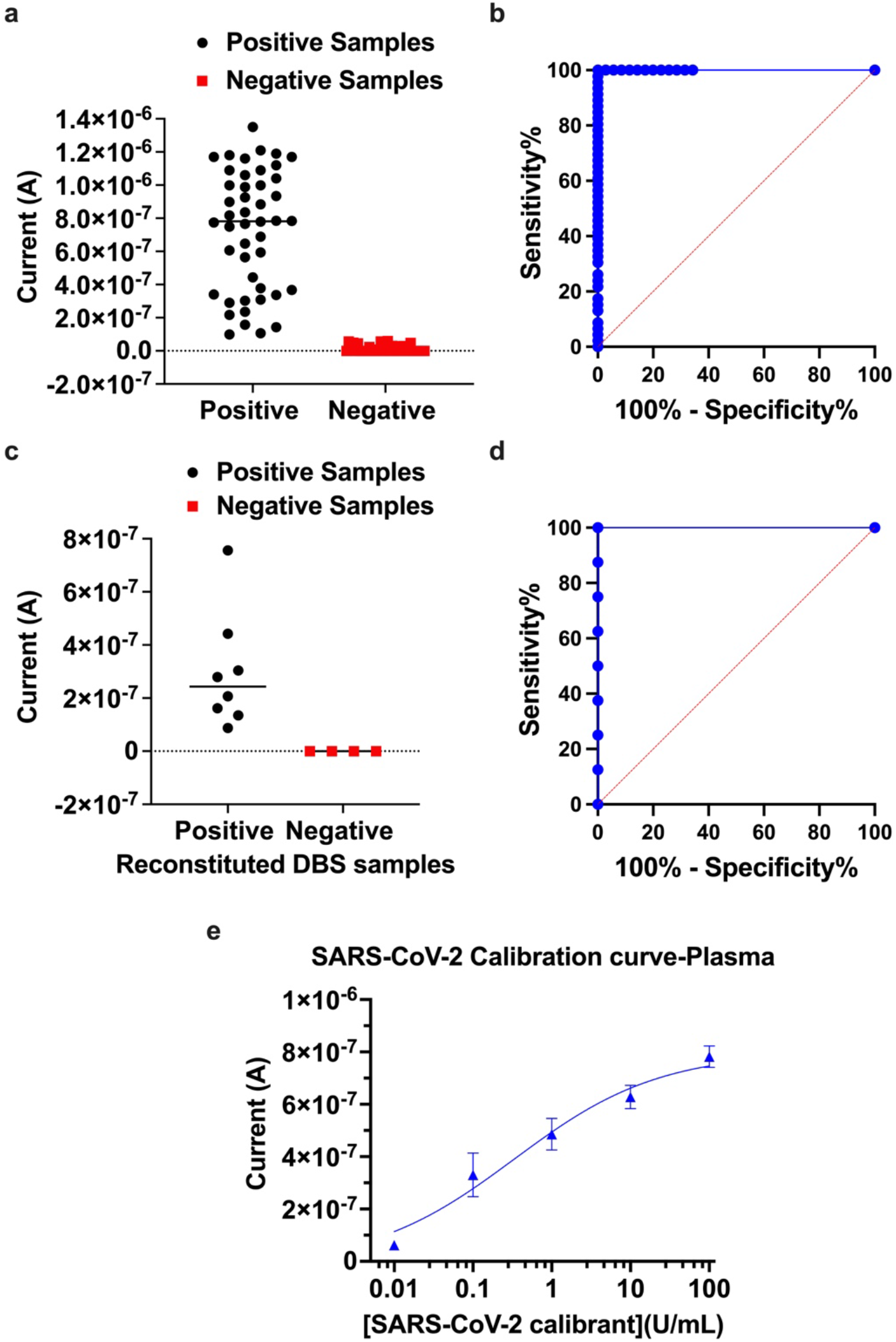
Clinical validation of EC-sensor for detection of SARS-CoV-2 IgG. Scattered graph (a) and ROC curve (b) for detection of 46 positive and 35 negative SARS-CoV-2 clinical plasma samples. Scattered graph (c) and ROC curve (d) for detection of 8 positive and 4 negative SARS-CoV-2 clinical Dried Blood Spot. (e) Calibration curve using NIBSC Anti-SARS-CoV-2 antibody diagnostic calibrant using pre-pandemic plasma sample. Error bars represent the s.d. of the mean; n = 3. Analysis was done using 4-Parameter Logistic (4PL) curve fitting.

The clinical utility of the quantitative detection of SARS-CoV-2 antibody using the EC sensor was further validated by generating a calibration curve using the NIBSC anti-SARS-CoV-2 antibody diagnostic calibrant. Initially, a calibration curve was run with 1% rHA in PBS (**Fig. S7b)**. However, after successful detection of the anti-SARS-CoV-2 antibody diagnostic calibrant in buffer, a pre-pandemic plasma sample without anti-SARS-CoV-2 IgG was used to run the calibration curve, which demonstrated a sensitivity of 0.01 U/mL (**Fig. 5e)**.

## 4. Conclusion and Discussion

In this study, we developed and validated a rapid multiplexed EC detection platform for monitoring COVID-19 and vaccination status by detecting SARS-CoV-2 antibodies with 100% sensitivity and specificity. The antifouling coating, we utilized increases the conductivity of the EC sensor and decreases nonspecific binding, which minimizes the EC background, hence increasing the sensitivity and selectivity of the assay. This EC sensor also can be used for rapid quantitative detection of SARS-CoV-2 IgG using just 1.5 µL of plasma sample within 10 min with 100% sensitivity and specificity. The potential of the EC-biosensor to perform assay with DBS was also demonstrated using clinical DBS samples with 100% sensitivity and specificity which can be used for serological surveillance utilizing remote sampling and shipment without refrigeration. In addition, validation of the quantitative capability of the EC-sensor was carried out by performing the calibration curve of a NIBSC SARS-CoV-2 calibrant in buffer and pre-pandemic plasma samples. Thus, these studies demonstrate the potential value of this antifouling EC-biosensor with its high sensitivity and selectivity for rapid quantitative detection of biomarkers in serologic assays that can be carried out in a multiplexed fashion, potentially at the point-of-care.

Antibody titers can remain stable over several months and extensive cohort studies in hospitalized patients show that IgG antibodies against viral proteins correlate with disease severity and outcome. In addition, rapid seroprevalence studies can differentiate reinfections versus breakthrough infections to better understand herd immunity and vaccine efficacy in the context of a pandemic, which is critical for the decision to reopen economies.^[6, 20]^ Hence, this type of serological assay could be used to provide critical information to respond to epidemics or pandemics, manage patient care and public health strategies, assess patient responses to vaccination, and help determine when and if boosters might be required.^[21]^

## Data Availability

All data produced in the present study are available upon reasonable request to the authors

## Statistical Analysis

ELISA reading is reported as absorbance (a.u.) of the mean of replicates and error bars represent the standard deviation (s.d.) of the mean; n = 2. For EC-biosensor studies, peak heights were calculated using Nova 1.11 software. Error bars represent mean ± s.d. for all EC-biosensor studies (sample sizes and statistical tests used are indicated in the Figure legends). All data was plotted, and statistical tests were performed using GraphPad Prism 8, and 4-Parameter Logistic (4PL) curve fitting was done for calibration curve analysis.

## Supporting Information

Supporting Information includes single step assay development, optimization of detection antibody concentration, screening of capture and detection antibody using ELISA, optimization of sample dilution, titration of high titer SARS-CoV-2 positive samples, BSA vs rHA characterization, and calibration curve of SARS-CoV-2 IgG.

## Data availability

The main data supporting the results in this study are available within the paper and its Supplementary Information. All raw and processed images/data generated in this work, including the representative images provided in the manuscript, are available from the corresponding authors on reasonable request.

## Acknowledgments

We acknowledge research funding from the Wyss Institute for Biologically Inspired Engineering at Harvard University and GBS Inc.

