## Supplementary Information for "Rapid quantitative electrochemical detection of SARS-CoV-2 antibodies in plasma and dried blood spot samples"

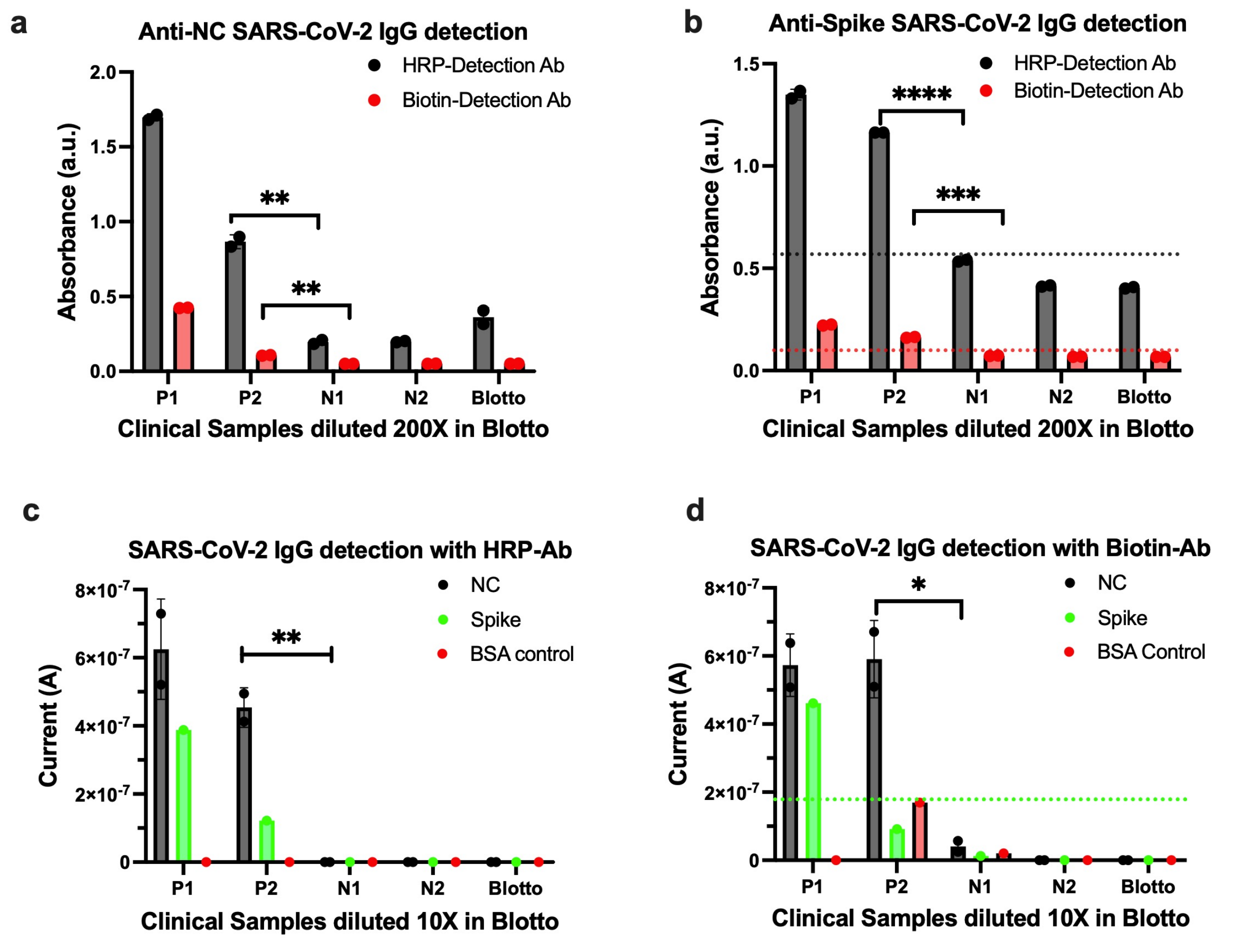


***Figure S1.*** *Assay development using biotin vs HRP labeled detection antibody. a) Anti-N SARS-CoV-2 detection with biotin vs HRP labeled detection antibody using ELISA; n=2. b) Anti-Spike SARS-CoV-2 detection with biotin vs HRP labeled detection antibody using ELISA; n=2. c) Anti-N and Spike SARS-CoV-2 detection with HRP labeled detection antibody using EC biosensor; n=3. d) Anti-N and Spike SARS-CoV-2 detection with biotin labeled detection antibody using EC biosensor; n=3. P= SARS-CoV-2 positive sample; N=SARS-CoV-2 negative sample; and Blotto=Negative Control. Green dotted line represents highest signal for negative sample/control. Statistical analysis was performed by unpaired t-test (ns P > 0.05; *P < 0.05); all two-tailed.*

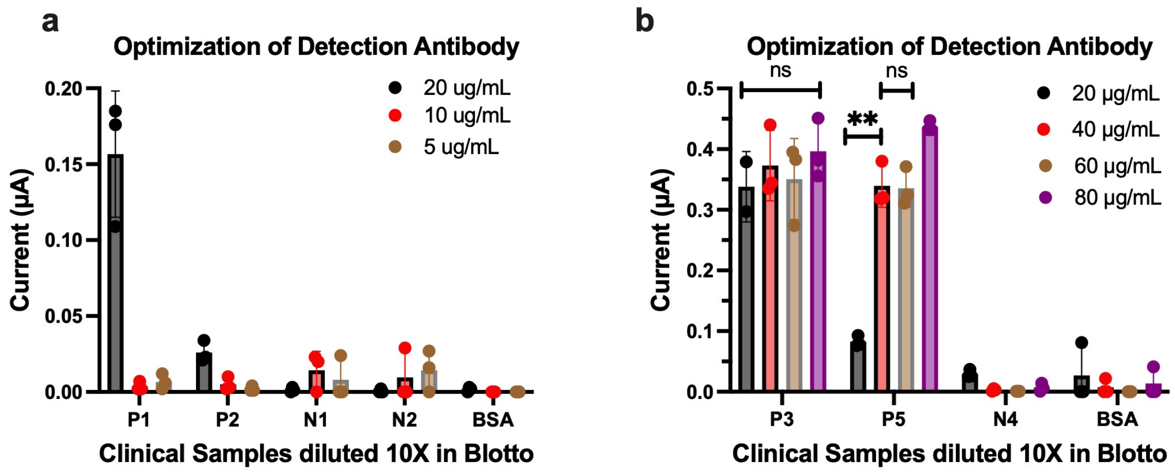


***Figure S2.*** *Optimization of anti-SARS-CoV-2 IgG detection antibody. a) Detection antibody optimization from 5-20 µg/mL. a) Detection antibody optimization from 20-80 µg/mL. n=3; P= SARS-CoV-2 positive sample; N=SARS-CoV-2 negative sample; and BSA=Negative Control. Statistical analysis was performed by unpaired t-test (ns P > 0.05; *P < 0.05); all two-tailed.*

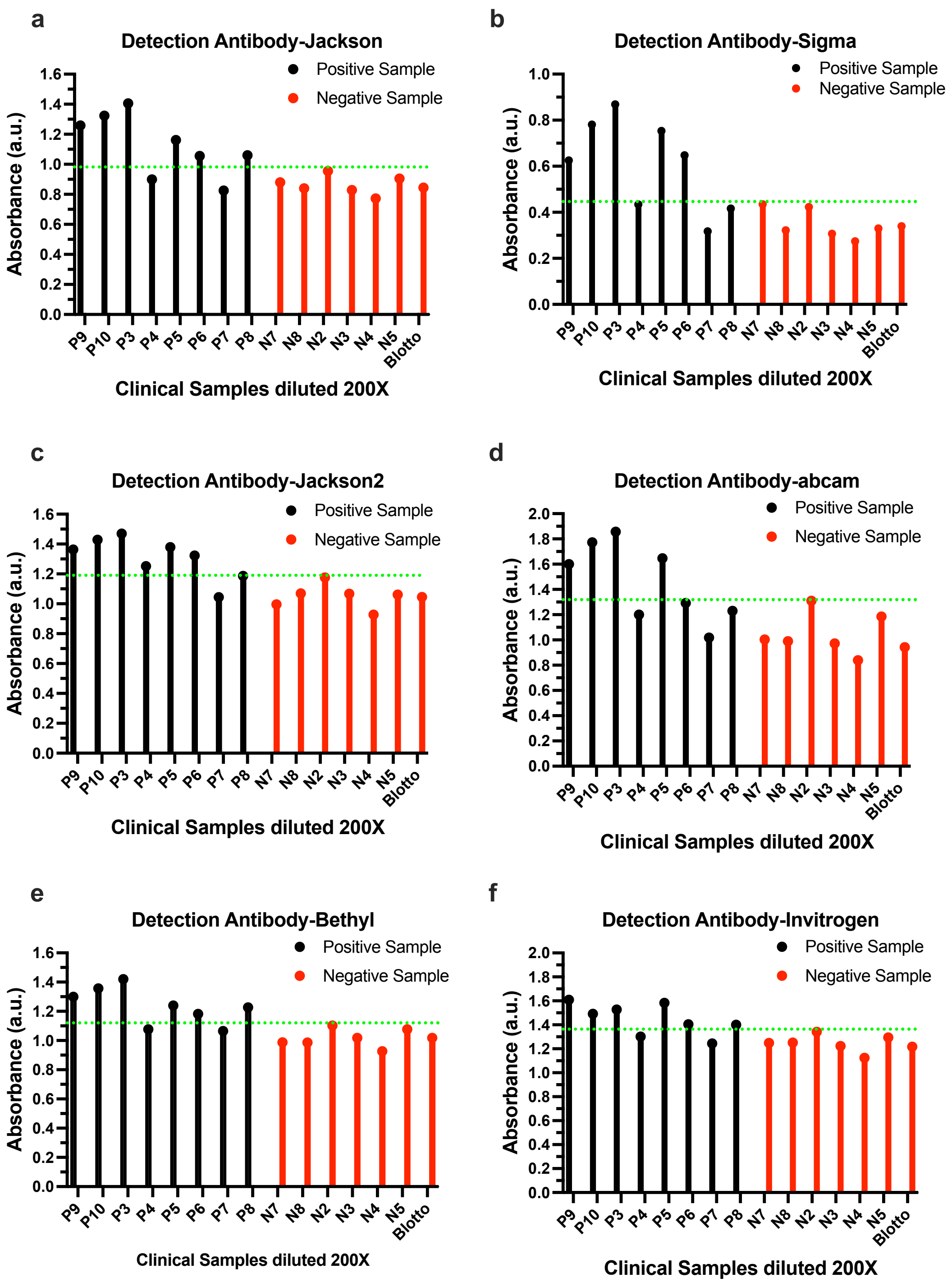


***Figure S3.*** *Screening of anti-human SARS-CoV-2 IgG detection antibody on ELISA from six commercial sources including Jackson (a), Sigma (b), Jackson (c), abcam (d), Bethyl (e), and Invitrogen (f). Error bars represent the s.d. of the mean; n=2; P= SARS-CoV-2 positive sample; N=SARS-CoV-2 negative sample; and Blotto=Negative Control. Green dotted line represents highest signal for negative sample/control.*

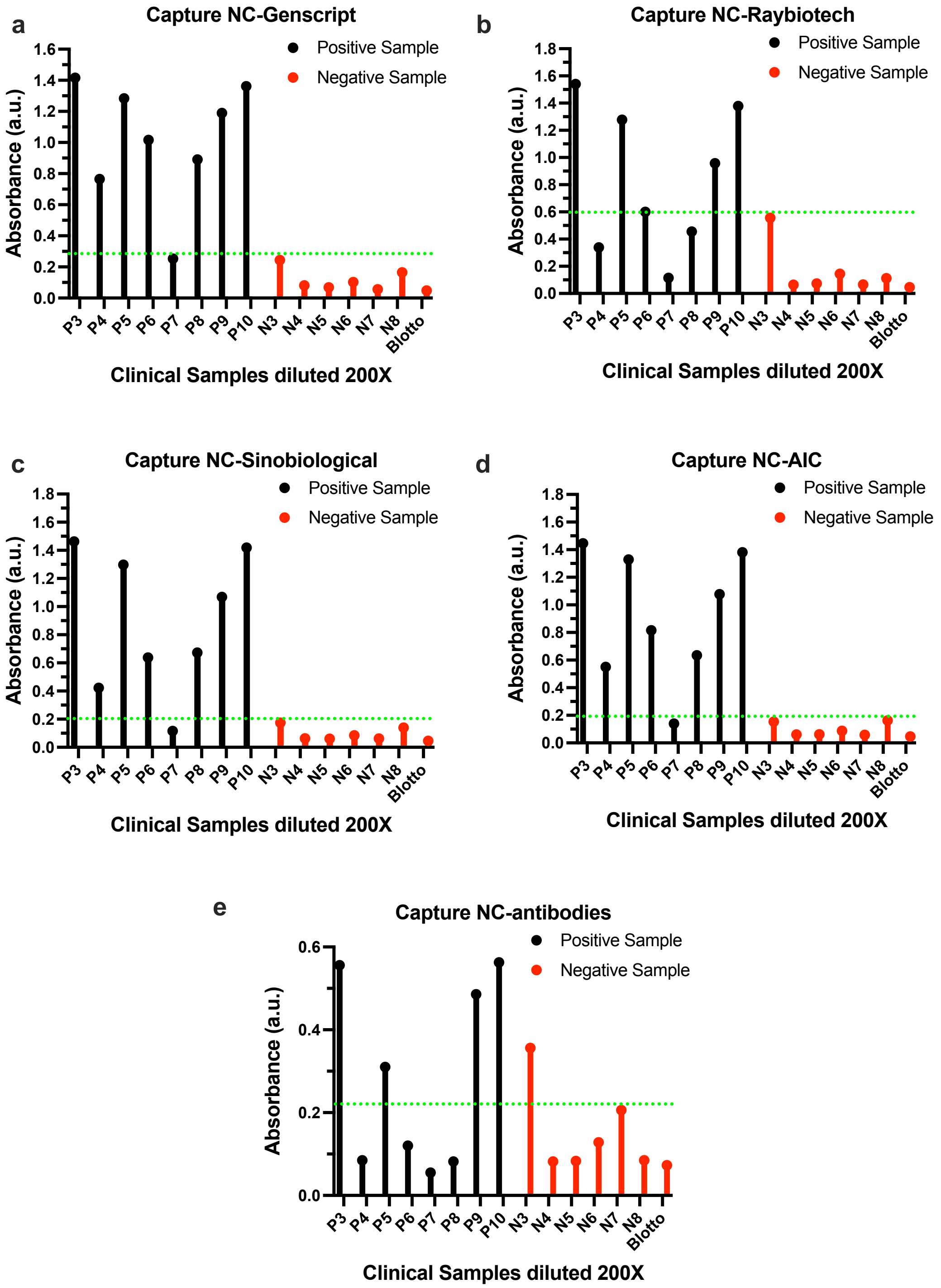


***Figure S4.*** *Screening of SARS-CoV-2 N capture protein on ELISA from five commercial sources including Genscript (a), RayBiotech (b), Sinobiological (c), AIC, and (d), Antibodies. Error bars represent the s.d. of the mean; n=2; P= SARS-CoV-2 positive sample; N=SARS-CoV-2 negative sample; and Blotto=Negative Control. Green dotted line represents highest signal for negative sample/control.*

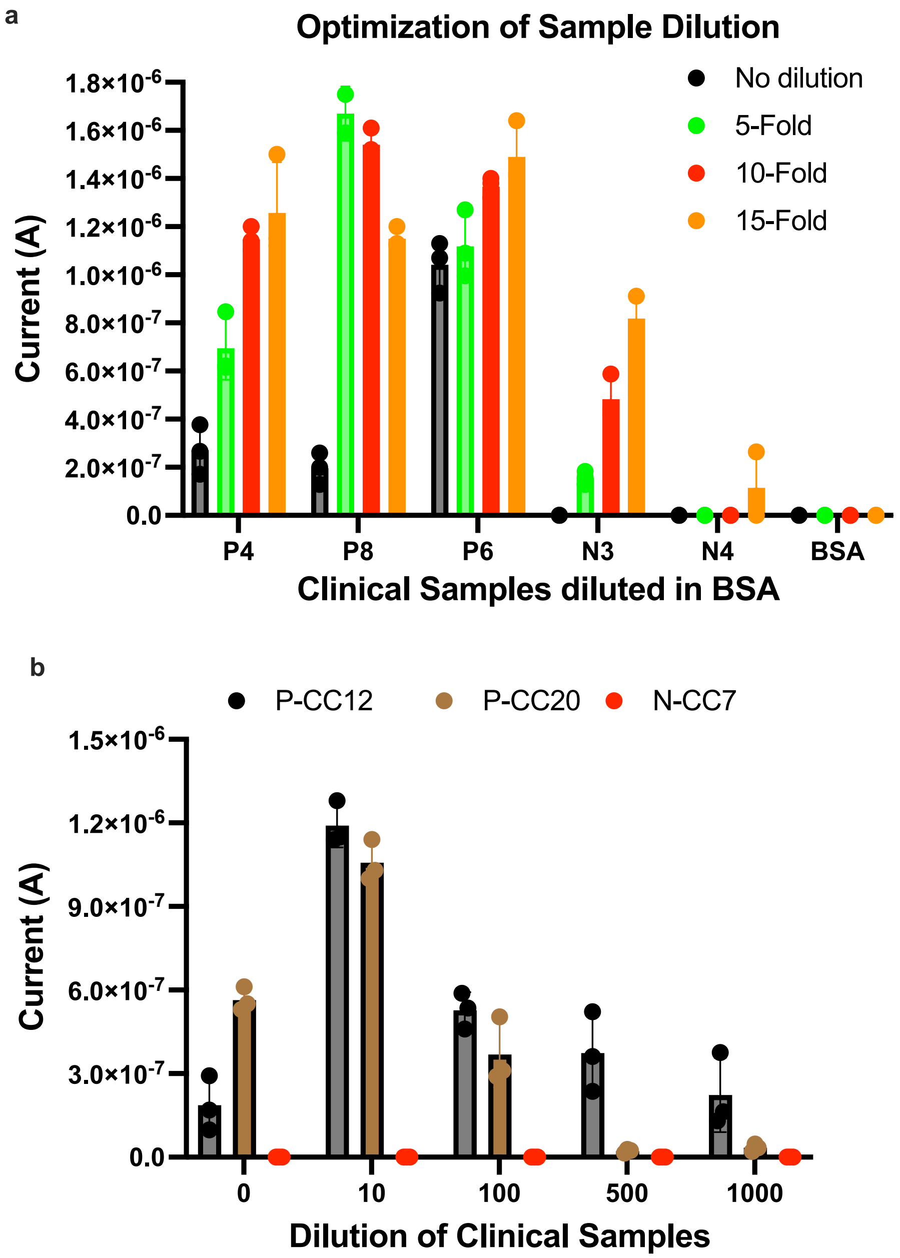


***Figure S5.*** *Sample dilution study for detection of SARS-CoV-2 in EC sensor. a) Optimization of sample dilution from 0-15-fold dilution for the detection of anti-SARS-CoV-2 IgG. b) Titration study of high titer anti-SARS-CoV-2 IgG clinical samples from 0-1000-fold dilution. Error bars represent the s.d. of the mean; n=3; P= SARS-CoV-2 positive sample; N=SARS-CoV-2 negative sample; and BSA=Negative Control.*

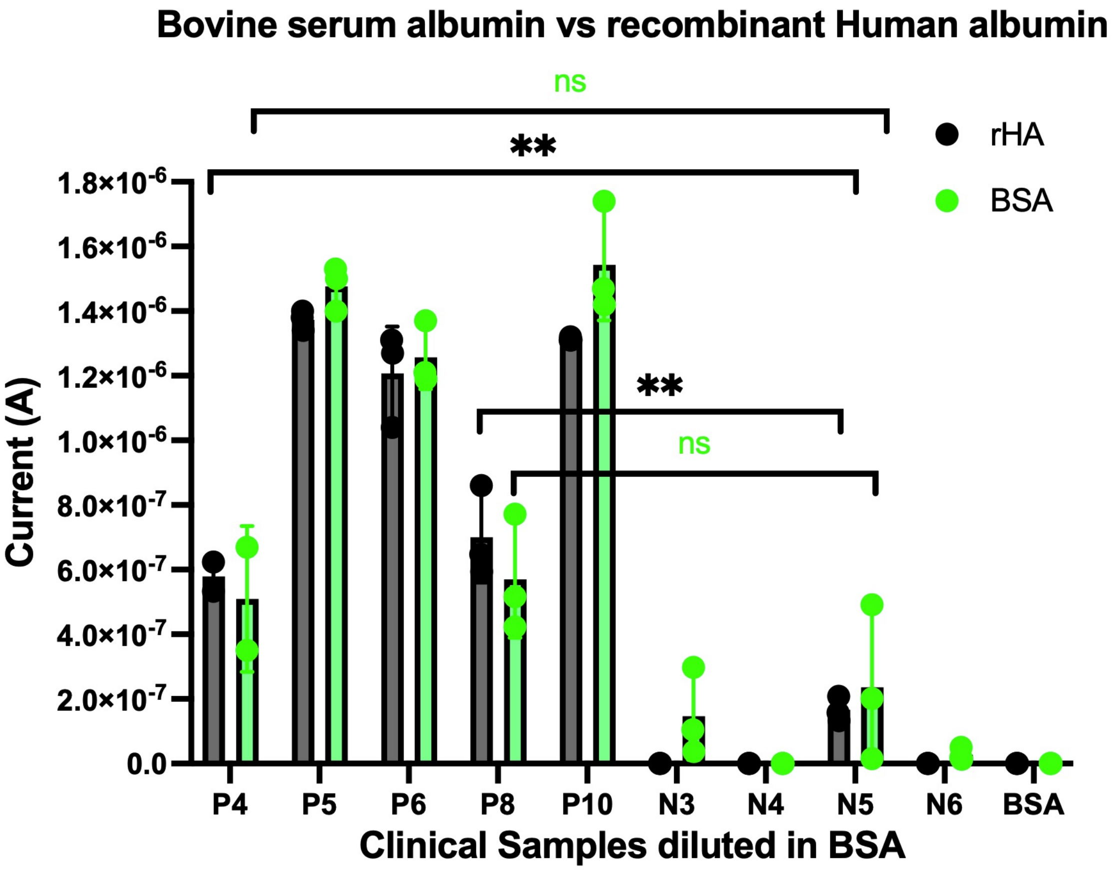


***Figure S6.*** *Comparison of BSA vs rHA for preparation of antifouling coating and preparation of buffer for the EC assay of SARS-CoV-2. Error bars represent the s.d. of the mean; n=3; P= SARS-CoV-2 positive sample; N=SARS-CoV-2 negative sample; and BSA=Negative Control. Statistical analysis was performed by unpaired t-test (ns P > 0.05; *P < 0.05; **P < 0.01; ***P < 0.001); all two-tailed.*

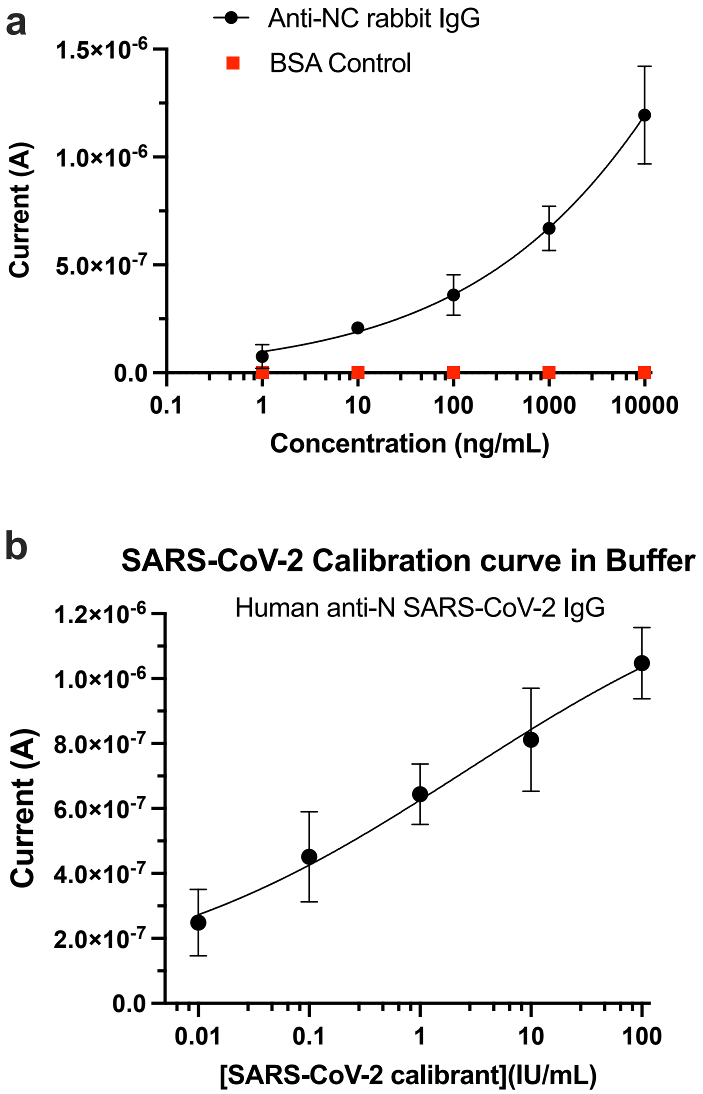


***Figure S7.*** *Calibration curve for detection of Anti- N SARS-CoV-2 antibody. a) Calibration curve of rabbit Anti-N SARS-CoV-2 antibody using buffer (1% BSA). b) Calibration curve using NIBSC human Anti-N SARS-CoV-2 antibody diagnostic calibrant using buffer (1% BSA). Error bars represent the s.d. of the mean; n = 3. Analysis was done using 4-Parameter Logistic (4PL) curve fitting.*
